# Interpretable multi-stream ensemble learning for radiographic pattern recognition

**DOI:** 10.1101/2021.08.09.21261788

**Authors:** José Raniery Ferreira, Diego Armando Cardona Cardenas

## Abstract

Chest radiography (CXR) remains an essential component to evaluate lung diseases. However, it is crucial nowadays to include computer-based tools to aid physicians in the early detection of chest abnormalities. Therefore, this work proposed deep ensemble models to improve the CXR evaluation, interpretability, and reproducibility. Five convolutional neural networks and six different processed image inputs yielded an AUC of 0.982. Furthermore, ensemble learning could produce more reliable outcomes as it did not consider the information of only one method. Moreover, the ensemble strategy balanced the most critical factors from each model to perform a more consistent classification. Finally, class activation and gradient propagation maps allowed locally visualizing CXR regions that most activate neurons from the trained models and explaining practically which areas of the CXR correlated to the model output.

## INTRODUCTION

Chest radiography (CXR) remains an essential component to evaluate patients with pathological suspicion, especially of lung diseases [1, 2]. However, it is essential to include computer-based tools to aid physicians to detect diseases early, as they can improve the accuracy and consistency of medical image diagnosis through computational support used as reference. Deep learning can analyze large volumes of images and automatically identify patterns accurately, including in CXR. However, most deep-learning methods were trained with single models, leading to limited prediction accuracy, even with optimum parameters [3].

One alternative to decrease this limitation is ensemble learning. This artificial intelligence paradigm designs a single model to combine different methods trained previously for the same task. The purpose of ensemble is to increase performance by combining the best properties from methods modeled with distinct configuration to achieve the highest efficiency and stability [4].

Furthermore, one huge limitation of general artificial neural networks is the black-box model produced by the training procedure. Thus, interpretable models are imperative for a robust computer-aided diagnostic tool. In this short paper, we propose deep ensemble models to improve the CXR -based evaluation, interpretability, and reproducibility.

## MATERIAL AND METHODS

This study used retrospectively anterior-posterior CXRs of patients suspected to have pneumonia [2]. The cohort contains a training set with 3,883 images with confirmed pneumonia and 1,349 normal individuals and a testing set with 234 normal images and 390 with pneumonia.

We used five traditional convolutional networks (ConvNets): DenseNet121, InceptionV3, MobileNet, VGG16, and Xception. All ConvNets were composed of the rectified linear unit and one composed of softmax function to classify the images. All models were previously trained in a preliminary work. To improve pattern recognition, we tested several image pre-processing methods based on image segmentation (such as adaptive thresholding and region growing), cropping (of the whole chest or lung-only regions, for instance), normalization (like based on Z-Score or Min-Max scaling), equalization (global or adaptive approaches), among others. We designed an extra artificial network for ensemble learning that concatenates the softmax probabilities of the single models. Data augmentation techniques (flip, scale, shift, and rotation) increased the number of images to reduce the possibility of overfitting and potentially improve performance.

## RESULTS

The ensemble of VGG16 obtained the highest overall efficiency, with an area under the receiver operating characteristic curve (AUC) of 0.973 (95% confidence interval - CI: 0.959–0.986), a sensitivity of 0.980 (CI: 0.964–0.992), and a specificity of 0.966 (CI: 0.940–0.987). Figure 1 presents some results indicating that ensemble learning may also produce more reliable outcomes as it would not consider the information of only one *observer*. The ensemble strategy balanced the most critical factors from each model to perform a more consistent classification.

**Figure 1:**
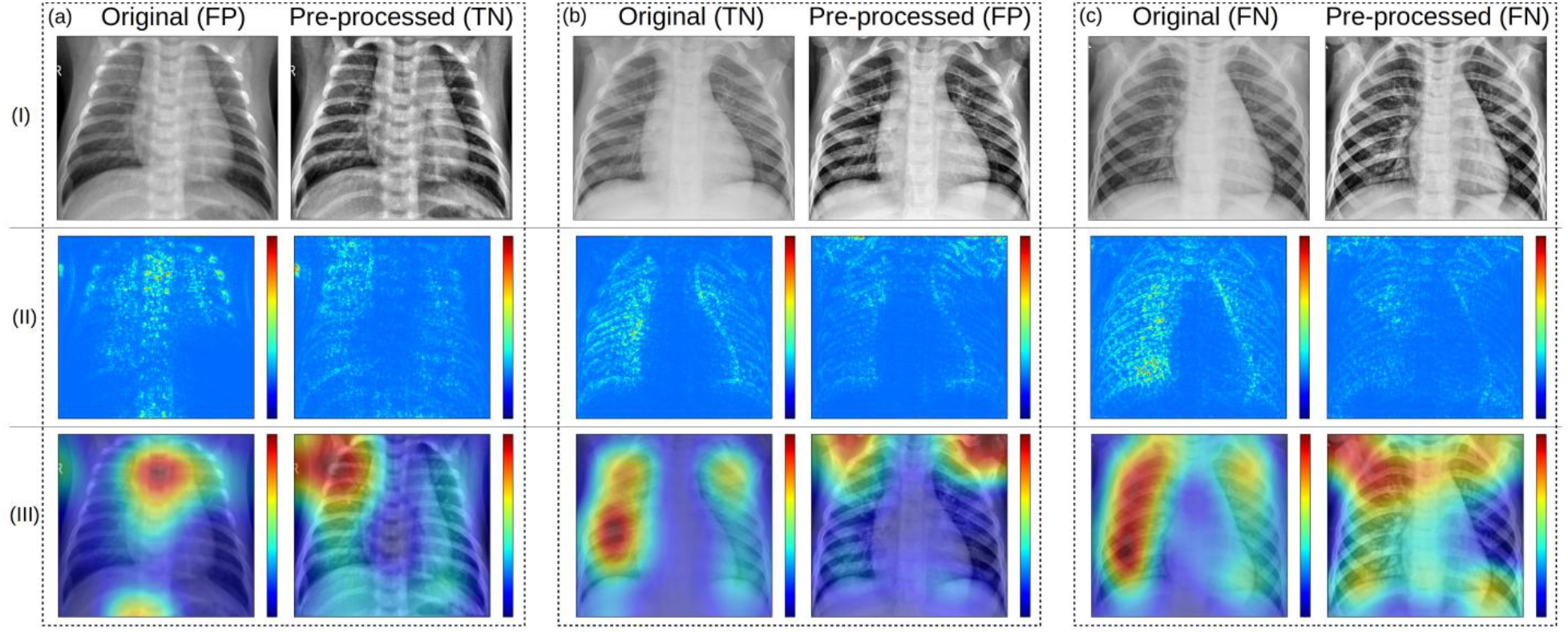
Examples of classification results with input image (row I) and corresponding guided- backpropagated gradients (row II) and CAM (row III). In (a), VGG16 correctly classified as a normal case using the pre-processed image as input but misclassified using the original image. In (b), VGG16 correctly classified as normal using the original image as input but misclassified using the pre-processed image. In (c), VGG16 misclassified as normal using both images as input, but the ensemble of them correctly classified the patient to have pneumonia. In the images, TN, FN, and FP correspond to true negative, false negative, and false positive, respectively.

Figure 1 also shows class activation maps (CAMs) and a gradient propagation approach to locally visualize the pixels of the CXR that most activate neurons from the trained models. These visual mechanisms highlighted discriminative parts from the image, allowing explaining practically which regions of the CXR correlated to the model output.

We also performed a comprehensive evaluation extending the ensemble exhaustively to the different ConvNets to find an optimum model. Combining all five ConvNets and six different image inputs yielded an AUC of 0.982, but no statistically significant difference was found.

## DISCUSSION

We herein introduced a multi-stream ensemble learning approach that could increase feature representation and identify intricate radiographic patterns, yielding higher AUCs than state-of-the-art single, limited models (*p* < 0.05) [2, 3, 5]. Moreover, the method automatically detects radiographic patterns, bringing significant benefits to the clinical routine at the beginning of care to prioritize abnormal exams for further reading from a specialist, ultimately optimizing examination time [6]. The models investigated could play an important role in the computer-aided detection of lung opacities. The correlation between radiography and chest abnormalities could accelerate the complementary exams’ adoption to confirm the biological nature of the abnormality.

It is becoming increasingly clear that ensemble learning provides cooperatively much more sophisticated results than would be practical from a single provider [ 4]. Ensemble learning has a good potential for external generalization due to the extra classification layers that take advantage of the different information from the ConvNets. They learn to weigh and associate the probabilities of different training strategies for the final classification.

Our main limitation in this study was the lack of an independent external dataset for generalization purposes. The most relevant CXR datasets publicly available are inconsistent due to the use of natural language processing for image labeling. This could lead to text-mining errors as labels may not accurately reproduce imaging patterns.

Finally, a tool implementing the proposed model could be useful for academic medicine. Resident students and specialists with low experience on CXR could improve image interpretation and opacity location skills using this approach. Moreover, it could promote a telemedicine tool to aid resource-limited places, where x-ray scanners are the only imaging option of health care.

## Data Availability

The dataset used in this work was made publicly available in Kermany D, Goldbaum M, Cai W, et al. Identifying medical diagnoses and treatable diseases by image-based deep learning. Cell. 2018;172:1122-1131.

